# Prevalence and predictors of hepatitis B virus infection among sellers and workers at West Africa’s largest market, Kejetia, Ghana

**DOI:** 10.1101/2025.09.18.25336118

**Authors:** De-Graft Kwaku Ofosu Boateng, Michael Agyemang Obeng, Joshua Kofi Attah, Abdul Latif Koney Shardow, Clinton Owusu Boateng, Daniel Kobina Okwan, Senam Yawa Nunamey Ahadzie, Pius Takyi, Godfred Yawson Scott, Akwasi Amponsah Abrampah, Felix Kusi Boakye, Patrick Oduro, Daniel Dzemedo Nouwati, Kofi Bonsu Amankwah, Reem Mahmoud Hassanin, Mohammed Mahi Adams, Abu Abudu Rahamani

**Affiliations:** Department of Pathology, Elbe Kliniken Stade-Buxtehude, Stade, Germany; Kumasi Centre for Collaborative Research in Tropical Medicine, Kwame Nkrumah University of Science and Technology, Kumasi, Ghana; Pathology Department, Komfo Anokye Teaching Hospital, Kumasi, Ghana; Department of Medical Diagnostics, Kwame Nkrumah University of Science and Technology, Kumasi, Ghana; Dawurampong Polyclinic, Ghana Health Service, Dawurampong, Ghana; Department of Anatomy, Kwame Nkrumah University of Science and Technology, Kumasi, Ghana; Maternal and Child Health Hospital, Ghana Health Service, Kumasi, Ghana; Public Health, Hamburg University of Applied Sciences, Hamburg, Germany; National Cardiothoracic Center, Korle Bu Teaching Hospital, Accra, Ghana; Department of Medical Laboratory Science, University for Development Studies, Tamale, Ghana

## Abstract

Hepatitis B virus (HBV) infection remains a major public health challenge in sub-Saharan Africa, including Ghana, despite the infant vaccination programs. Limited data exist on HBV prevalence and predictors in informal sector populations, who may face unique occupational and behavioural exposures. This study assessed the prevalence and predictors of HBV infection among sellers and workers at Kejetia Market, Ghana’s largest commercial hub.

A cross-sectional study was conducted among 489 adult market workers from 4^th^ December, 2024 to 25^th^ January, 2025. Participants were selected through stratified random sampling across occupational groups. Data on sociodemographic, occupational, and behavioural factors were collected using structured questionnaires. On-site testing for hepatitis B surface antigen (HBsAg) was performed using the Hightop One Step Rapid Test kit. Bivariate and multivariate logistic regression analyses were used to identify independent predictors of HBV infection. P < 0.05 was considered statistically significant.

The overall prevalence of HBV infection was 7.36% (36/489), consistent with intermediate-to-high endemicity. Multivariate analysis identified three independent predictors of HBV infection. Female gender (aOR = 0.455, 95% CI: 0.221–0.937; p = 0.033) and absence of tattoos (aOR = 0.283, 95% CI: 0.110–0.730; p = 0.009) were associated with lower risk of HBV infection, while unvaccinated individuals had 3.37-fold increased odds of getting the infection (95% CI: 1.395– 8.142; p = 0.007). HBV prevalence declined progressively with increasing vaccine doses, from 9.2% in unvaccinated individuals to 2.3% among those who had completed three or more doses.

HBV infection is common among Kejetia market workers, with prevalence exceeding both continental and global estimates. Gender, tattooing, and vaccination status were significant predictors of infection. Strengthening adult vaccination programs, promoting safe tattooing practices, and implementing male-focused screening and prevention interventions are critical to reducing HBV burden and achieving Ghana’s contribution to the WHO goal of eliminating HBV by 2030.

## Introduction

Hepatitis B virus (HBV) infection affects 296 million people worldwide, accounting for approximately 1.1 million deaths annually, mainly due to cirrhosis and hepatocellular carcinoma[1,2]. This burden is disproportionately concentrated in the Western Pacific and sub-Saharan Africa (SSA)[3]. Ghana illustrates this with an overall HBV prevalence of 8.36% among adults, underscoring the country’s high endemic status by World Health Organization (WHO) categorization[4].

Preventive efforts in Ghana have mostly targeted infants, with the introduction of a pentavalent vaccine in 2002 that combines HBV with other essential infectious disease vaccines into the national immunization program[5]. This drive can be attributed to the significant reduction to 2% overall HBV prevalence among children in Ghana and Senegal[6]. Despite this effort, HBV remains a significant public health issue in Ghana among the adult populace[4,7,8].

Several factors influence HBV infection in Ghana. Body modifications (scarification, tattooing, piercing, circumcision) are critical infection pathways as well as exposure to infected blood and saliva, reuse and sharing of sharp objects and personal items, sexual activities and lack of vaccination[6,9,10].

Occupational factors play an essential role in HBV infection and transmission risk among specific population groups. Previous studies have shown that specific occupational groups such as barbers and long-distance drivers exhibit higher HBV prevalence (14.40%), suggesting the inherent risk of infection associated with certain professions and their occupational environment[4]. These findings suggest that certain workplace exposures, safety practices and access to preventive measures can influence the infection risk [11]. It is thus relevant to understand the role occupational and behavioural factors play in HBV prevalence.

Previous studies on HBV in Ghana have focused on pregnant women, blood donors and the clinical settings[4,12]. However, there is a scarcity of data focusing on prevalence and predictors of HBV infection within the non-clinical setting especially among informal sector workers in high-traffic commercial hubs like the Kejetia market.

Kejetia market is one of the largest and busiest commercial hubs in West Africa with over 20,000 vendors. These workers may be at increased risk of HBV infection due to their work environment, including factors such as close interpersonal interactions and frequent exchange of goods, frequent exposure to sharp objects among specific occupations, and potential close contact with infected individuals[13,14]. The market’s pivotal role in the regional economy suggests that health interventions in this target population could possess sweeping benefits.

This study aimed to assess the prevalence and predictors of HBV infection among sellers and workers at the Kejetia market. Specifically, we sought to identify the sociodemographic and behavioural factors that may predispose this population to HBV infection.

This research addresses critical gaps in understanding HBV dynamics in the informal work settings. Findings will inform context-specific prevention strategies, resource allocation and policy guiding decisions on the National Viral Hepatitis Control Programme (NVHCP), contributing to achieving the Sustainable Development Goal (SDG) of reducing Hepatitis B mortality by 65% by 2030[15]. Given the prevalence of similar commercial environments throughout sub-Saharan Africa, insights from this study may have broad regional relevance for HBV prevention efforts across the continent.

## Materials and Methods

### Study design

A cross-sectional study design was employed to assess the prevalence and determinants of HBV infection among sellers and workers at Kejetia market, Kumasi, Ghana from 4^th^ December, 2024 to 25^th^ January, 2025. This was done as it allowed for the simultaneous assessment of HBV status and its associated risk factors at a specific point in time within the target population.

### Study setting and population

The study was conducted at the Kejetia market, Kumasi, one of the largest commercial hubs in the West Africa. With over 8,000 stores and stalls and about 20,000 vendors, the market hosts diverse range of occupational groups engaged in daily activities[13,14]. The target population comprised adult sellers and workers (≥18 years) across various occupational categories, including butchers, fishmongers, tailors, head porters, drivers, food vendors, and general goods storekeepers. These occupational groups were selected due to their potential differential exposure to HBV risk factors, including occupational injuries, close interpersonal interactions, personal hygiene practices, and varying levels of healthcare access.

### Sampling technique

A total of 489 participants were recruited using a stratified random sampling technique to ensure adequate representation of different occupational groups within Kejetia market. The market population was first categorized into predefined strata based on work type. Within each occupational stratum, participants were randomly selected using a systematic random sampling approach where every n^th^ individual was selected after identifying a random starting point.

The minimum sample size was calculated to be 377 participants using the Raosoft sample size calculator at 95% confidence level, a 5% margin of error, an estimated population size of 20,000 and a response distribution of 50%[16]. The total sample size was increased to 489 to improve statistical power.

### Ethical consideration

Ethical approval was obtained from the Committee on Human Research, Publication and Ethics (CHRPE) of the School of Medical Sciences, KNUST with approval number CHRPE/AP 180/24. Kejetia management gave site approval to conduct the study at the Market. Written informed consent was obtained from all participants after explaining the purpose, procedures, benefits, and potential risks of the study in their preferred language.

Confidentiality was maintained by using unique identification codes in place of personal identifiers on all study documents. Participants found to be HBsAg-positive were counselled and referred to appropriate healthcare facilities for further evaluation and management.

### Inclusion and exclusion criteria

The study participants were adult sellers and workers (≥18 years) who gave written informed consent and had worked at the site for a minimum of six months. Persons who were incapable of providing informed consent due to a mental health disorder were excluded.

### Data collection

A well-structured questionnaire was pretested, validated and administered through face-to-face interviews with the participants to collect data on their sociodemographic, occupational, HBV vaccination status and behavioural factors.

Data collection was conducted from 4^th^ December, 2024 to 25^th^ January, 2025 by trained research assistants proficient in local languages. Before commencement of the study, all researchers underwent training and retraining prior to the pretesting of the questionnaire. After each day of data collection, a cadre of investigators meticulously reviewed the data obtained, checking for inconsistencies and omissions. The questionnaire used in this study is presented in the supplementary materials (S1 Appendix).

### HBsAg testing

Following the interviews, on-site testing was conducted using finger-prick blood samples. The fingertip of each participant was thoroughly disinfected with 70% ethanol and allowed to dry completely before performing the finger prick using disposable sterile lancets. This allowed for collection of whole blood samples for immediate testing.

Serological testing for HBV surface antigen (HBsAg) was performed on-site using the Hightop HBV One Step Rapid Test (Hightop Biotech Co., Ltd., Qingdao, China) according to the manufacturer’s instructions. The whole blood sample obtained from finger prick was applied directly to the test device, and buffer solution was added as specified in the manufacturer’s protocol with results read within 15-20 minutes. All test sample results were then confirmed by a second person for positivity or negativity. Quality control measures were implemented throughout the testing procedures, including the use of positive and negative controls with each new batch of test kits to ensure accuracy and reliability of results.

The Hightop One Step Rapid Test was used for HBsAg detection because of its practicality in large field-based studies where laboratory infrastructure is limited. HBsAg rapid diagnostic tests have shown high diagnostic accuracy in meta-analysis, with pooled sensitivity around 90% and specificity around 99.5% compared to immunoassays[17]. Its ability to provide immediate results was crucial in this study, as it allowed on-site counselling, referral of positive cases, and minimized loss to follow-up.

### Data analysis

Data collected were entered onto Microsoft Excel sheet and exported to IBM SPSS Version 26.0 and GraphPad Prism Version 8.0 for analysis. Categorical data were presented as frequencies (proportions), while continuous variables were summarized using means with standard deviations or medians with interquartile ranges. The overall prevalence of HBV infection and occupation-specific prevalence rates were calculated. Chi-square tests were used to examine associations between independent variables and hepatitis b infection status. Logistic regression analysis was performed to identify independent predictors of hepatitis B infection. Crude odds ratios (cOR) and adjusted odds ratios (aOR) with 95% confidence intervals (CI) were reported. A p-value < 0.05 was considered statistically significant.

## Results

### Sociodemographic and behavioural factors associated with hepatitis B infection

The age group with the highest frequency was ≤ 30 years 159(32.5%). Most of the participants were female 279(57.1%) and were married 291(59.5%). Majority of them were Christians 334(68.3%), had basic education 240(49.1%), and have stayed in urban areas for greater part of their lives 358(73.2%). Also, more than half 307(62.8%) of them had not received any education on Hepatitis B infection. Majority of them 361(73.4%) had never contracted STIs and most of them 369(75.5%) had their first sexual intercourse ≥18 years old. Over two-third of them, 437(89.4%) had never received blood transfusion and did not have tattoos 451(92.2%). Additionally, most of them did not have close person living with hepatitis B 420(85.9%), and had had their fingernails/toenails clipped by commercial nail clippers before 349(71.4%) and majority of them had multiple sexual partners 351(71.8%). Most 306(62.6%) of them had not taken any shot of the hepatitis B vaccine (**Table 1**).

**Table 1.**
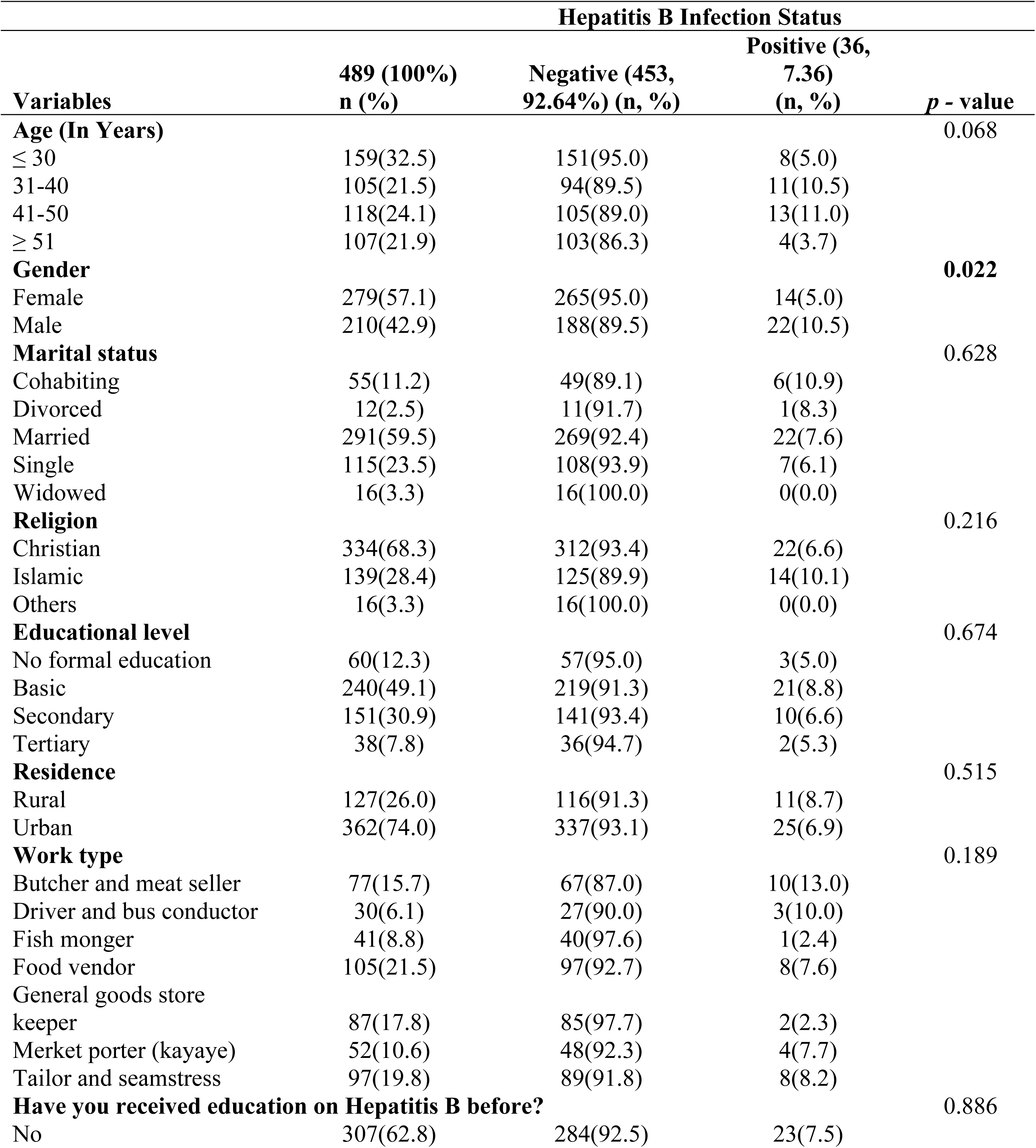
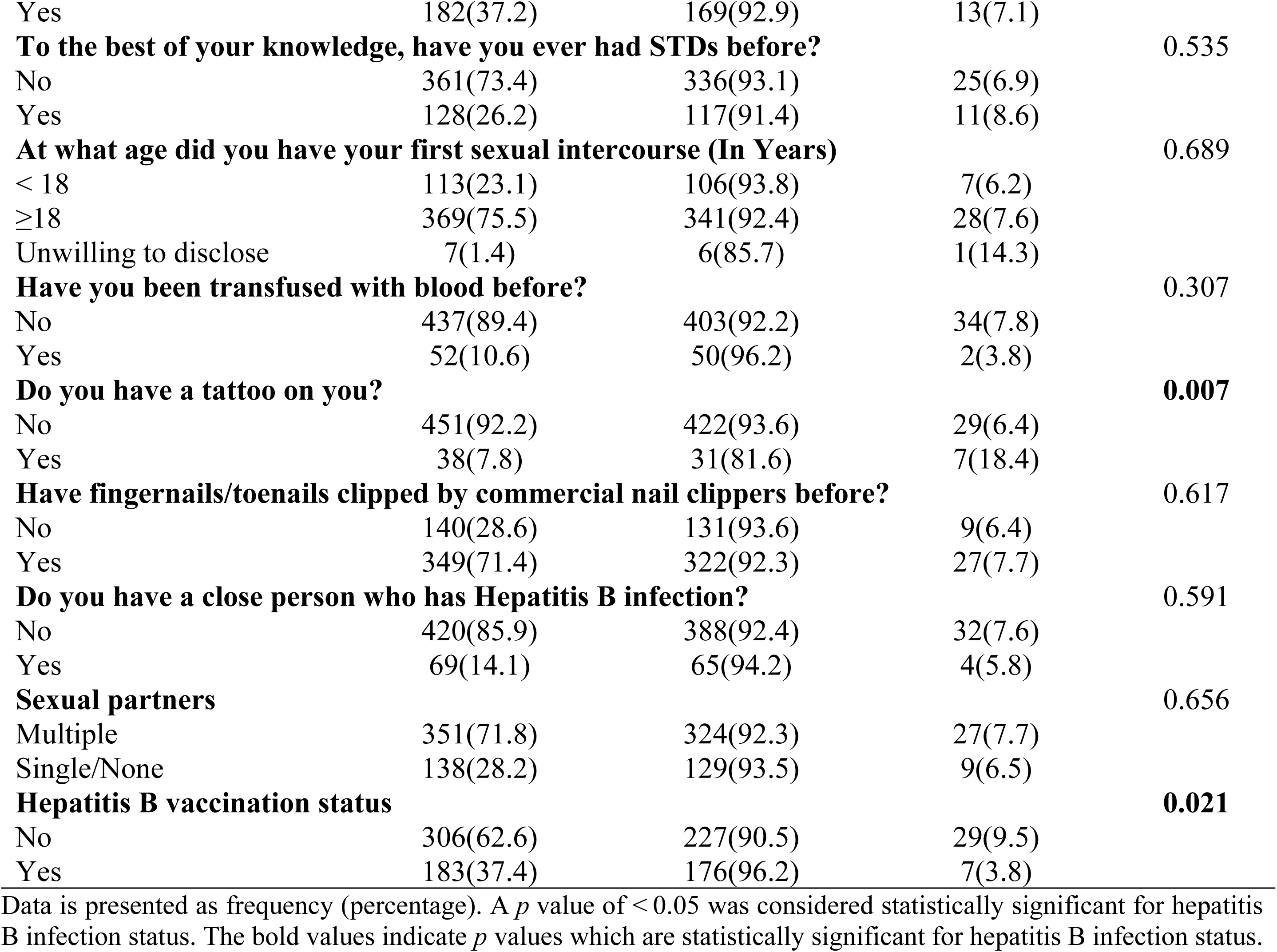
Sociodemographic and Behavioural Factors Associated with Hepatitis B Infection.

| Variables | Hepatitis B Infection Status |  |  | *p* - value |
| --- | --- | --- | --- | --- |
|  | 489 (100%)<br>n (%) | Negative (453,<br>92.64%) (n, %) | Positive (36,<br>7.36)<br>(n, %) |  |
| **Age (In Years)** |  |  |  | 0.068 |
| ≤ 30 | 159(32.5) | 151(95.0) | 8(5.0) |  |
| 31-40 | 105(21.5) | 94(89.5) | 11(10.5) |  |
| 41-50 | 118(24.1) | 105(89.0) | 13(11.0) |  |
| ≥ 51 | 107(21.9) | 103(86.3) | 4(3.7) |  |
| **Gender** |  |  |  | **0.022** |
| Female | 279(57.1) | 265(95.0) | 14(5.0) |  |
| Male | 210(42.9) | 188(89.5) | 22(10.5) |  |
| **Marital status** |  |  |  | 0.628 |
| Cohabiting | 55(11.2) | 49(89.1) | 6(10.9) |  |
| Divorced | 12(2.5) | 11(91.7) | 1(8.3) |  |
| Married | 291(59.5) | 269(92.4) | 22(7.6) |  |
| Single | 115(23.5) | 108(93.9) | 7(6.1) |  |
| Widowed | 16(3.3) | 16(100.0) | 0(0.0) |  |
| **Religion** |  |  |  | 0.216 |
| Christian | 334(68.3) | 312(93.4) | 22(6.6) |  |
| Islamic | 139(28.4) | 125(89.9) | 14(10.1) |  |
| Others | 16(3.3) | 16(100.0) | 0(0.0) |  |
| **Educational level** |  |  |  | 0.674 |
| No formal education | 60(12.3) | 57(95.0) | 3(5.0) |  |
| Basic | 240(49.1) | 219(91.3) | 21(8.8) |  |
| Secondary | 151(30.9) | 141(93.4) | 10(6.6) |  |
| Tertiary | 38(7.8) | 36(94.7) | 2(5.3) |  |
| **Residence** |  |  |  | 0.515 |
| Rural | 127(26.0) | 116(91.3) | 11(8.7) |  |
| Urban | 362(74.0) | 337(93.1) | 25(6.9) |  |
| **Work type** |  |  |  | 0.189 |
| Butcher and meat seller | 77(15.7) | 67(87.0) | 10(13.0) |  |
| Driver and bus conductor | 30(6.1) | 27(90.0) | 3(10.0) |  |
| Fish monger | 41(8.8) | 40(97.6) | 1(2.4) |  |
| Food vendor | 105(21.5) | 97(92.7) | 8(7.6) |  |
| General goods store<br>keeper | 87(17.8) | 85(97.7) | 2(2.3) |  |
| Market porter (kayaye) | 52(10.6) | 48(92.3) | 4(7.7) |  |
| Tailor and seamstress | 97(19.8) | 89(91.8) | 8(8.2) |  |
| **Have you received education on Hepatitis B before?** |  |  |  | 0.886 |
| No | 307(62.8) | 284(92.5) | 23(7.5) |  |
| Yes | 182(37.2) | 169(92.9) | 13(7.1) |  |
| **To the best of your knowledge, have you ever had STDs before?** |  |  |  | 0.535 |
| No | 361(73.4) | 336(93.1) | 25(6.9) |  |
| Yes | 128(26.2) | 117(91.4) | 11(8.6) |  |
| **At what age did you have your first sexual intercourse (In Years)** |  |  |  | 0.689 |
| < 18 | 113(23.1) | 106(93.8) | 7(6.2) |  |
| ≥18 | 369(75.5) | 341(92.4) | 28(7.6) |  |
| Unwilling to disclose | 7(1.4) | 6(85.7) | 1(14.3) |  |
| **Have you been transfused with blood before?** |  |  |  | 0.307 |
| No | 437(89.4) | 403(92.2) | 34(7.8) |  |
| Yes | 52(10.6) | 50(96.2) | 2(3.8) |  |
| **Do you have a tattoo on you?** |  |  |  | **0.007** |
| No | 451(92.2) | 422(93.6) | 29(6.4) |  |
| Yes | 38(7.8) | 31(81.6) | 7(18.4) |  |
| **Have fingernails/toenails clipped by commercial nail clippers before?** |  |  |  | 0.617 |
| No | 140(28.6) | 131(93.6) | 9(6.4) |  |
| Yes | 349(71.4) | 322(92.3) | 27(7.7) |  |
| **Do you have a close person who has Hepatitis B infection?** |  |  |  | 0.591 |
| No | 420(85.9) | 388(92.4) | 32(7.6) |  |
| Yes | 69(14.1) | 65(94.2) | 4(5.8) |  |
| **Sexual partners** |  |  |  | 0.656 |
| Multiple | 351(71.8) | 324(92.3) | 27(7.7) |  |
| Single/None | 138(28.2) | 129(93.5) | 9(6.5) |  |
| **Hepatitis B vaccination status** |  |  |  | **0.021** |
| No | 306(62.6) | 227(90.5) | 29(9.5) |  |
| Yes | 183(37.4) | 176(96.2) | 7(3.8) |  |

Data is presented as frequency (percentage). A $p$ value of $< 0.05$ was considered statistically significant for hepatitis B infection status. The bold values indicate $p$ values which are statistically significant for hepatitis B infection status.

There was significant association between gender (*p***=**0.0220), presence of tattoos (*p***=**0.007), hepatitis B vaccination status (*p***=**0.021) and Hepatitis B infection. However, age (*p*=0.068), marital status (*p*=0.628), religion (*p*=0.216), educational level (*p*=0.674), residence (*p*=0.515), work type (*p*=0.189), awareness of hepatitis B (*p*=0.886), history of sexually transmitted diseases (STDs) (*p*=0.535), age at first sexual intercourse (*p*=0.689), history of blood transfusion (*p*=0.307), experiences with fingernail or toenail clipping (*p*=0.617), having close person with hepatitis B infection (*p*=0.591), sexual partners (*p*=0.656), and hepatitis B vaccination status (*p*=0.021) were not significantly associated with hepatitis B infection status (**Table 1**).

### Hepatitis B infection prevalence and its distribution per vaccine shot among study participants

Figure 1A illustrates the percentage of individuals who tested positive for Hepatitis B infection in relation to the number of Hepatitis B vaccine shots received. Figure 1B also shows the overall prevalence of Hepatitis B infection.

**Figure 1.**
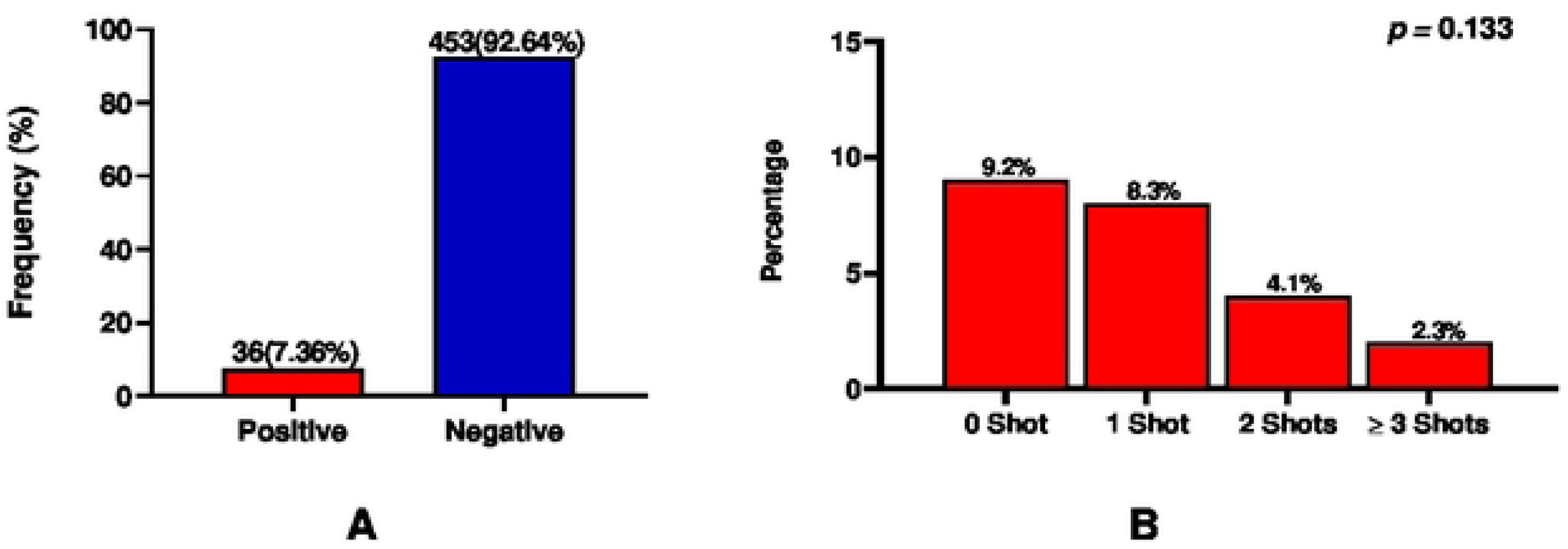
Hepatitis B Infection Prevalence and Its Distribution per Vaccine Shot Among Study Participants.

Out of the 489 participants recruited for the study, 36(7.36%) of them tested positive for Hepatitis B infection (**Fig 1B**).

The highest proportion of Hepatitis B positive cases was observed among individuals who had not received any vaccine doses, accounting for 9.2% HBV infection prevalence of the group. Among those who had received one dose, 8.3% tested positive. The percentage of positive cases further declined to 4.1% in those who had received two doses, and was lowest (2.3%) among individuals who had received three or more doses (**Fig 1B**).

### Predictors of hepatitis B infection

In the univariate logistic regression model, gender, presence of tattoo, and vaccination status were the predictors of hepatitis B infection status. In the multivariate logistic regression model, female (aOR = 0.455, 95% CI [0.221-0.937], *p* =0.033), not having tattoo (aOR = 0.283, 95% CI [0.110-0.730], *p* =0.009), not taking the vaccine (aOR = 3.371, 95% CI [1.395-8.142], *p* =0.007) were the independent predictor of hepatitis B infection (**Table 2**).

**Table 2.** Predictors of Hepatitis B infection.

| Variables | cOR | *p* - value | aOR | *p* - value |
| --- | --- | --- | --- | --- |
| **Sex** |  |  |  |  |
| Female | 0.451(0.225-0.905) | **0.025** | 0.455(0.221-0.937) | **0.033** |
| Male | 1 |  |  |  |
| **Do you have a tattoo on you?** |  |  |  |  |
| No | 0.304(0.123-0.750) | **0.010** | 0.283(0.110-0.730) | **0.009** |
| Yes | 1 |  |  |  |
| **Vaccination status** |  |  |  |  |
| No | 0.380(0.163-0.886) | **0.025** | 3.371(1.395-8.142) | **0.007** |
| Yes | 1 |  |  |  |

1.00, reference. Binary logistic regression analysis was performed to obtain odds ratios. A $p$ value of $<0.05$ was considered statistically significant for hepatitis B infection status. The bold values indicate $p$ values which are statistically significant for hepatitis B infection status. Abbreviations: aOR, adjusted odds ratio; CI, confidence interval; cOR, crude odds ratio.

## Discussion

This study examined the prevalence and predictors of HBV infection among sellers and workers at the Kejetia market, Ghana’s largest commercial hub. The overall prevalence of HBV infection in the study population was 7.36% (Fig 1A). This prevalence falls within the intermediate-to-high endemic range as defined by WHO [18]. This highlights the fact that HBV infection is still a major public health concern in Ghana. The finding is consistent with 7.5% HBV prevalence reported among blood donors in Volta region of Ghana [19], 7.2% prevalence among adults in Northern Ghana[20] and 7.88% in Southern Ethiopia[21]. The current finding is slightly lower than the pooled HBV prevalence of 8.36% among adults in Ghana by Abesig *et al.* [4] and crude prevalence of 8.48% by Nartey *et al.* [22], suggesting that the Kejetia market population mirrors the broader regional epidemiological pattern. In contrast, the current prevalence is significantly lower than the prevalence of 15.6 % by Helebge *et al.* among market women in the northern part of the country[23]. It is important to note that this study was done about five years prior to the current study. Today, increased education and awareness leading to preventive practices such as vaccine uptake against HBV infection could have contributed to the observed comparatively reduced prevalence. Nonetheless, this sharp contrast in the prevalence rate could be due to differences in geographical locations and lifestyles as well. It is recommended for further studies to explore this occurrence. Still, the finding is higher than prevalence studies reported in other parts of the country such as 4.4% among pregnant women in Eastern Ghana [24], 5.9% among healthcare workers in Southern Ghana [7] and 2.4% among pregnant women in Volta region of Ghana[25]. The HBV prevalence in the current study is comparatively higher than that of the 6.9% for blood donors and 5.9% for pregnant women reported for Africa in current systematic review and meta-analysis[26,27]. Again, the current prevalence is more than double the global HBV prevalence (3.2%) recently reported [28], indicating a high community burden in Kejetia adult population. This emphasizes that more efforts are required in Ghana to achieve the World Health Organization’s goal to eliminate HBV infection by 2030[29].

Being female appeared to be protective against HBV infection, as females were almost 0.5 times as likely to be infected with Hepatitis B compared to males (Table 2.0). This is consistent with prior reports from other studies, where men often exhibit higher HBsAg prevalence compared to women[30–33]. Possible explanations include gender differences in health-seeking behaviour, occupational exposure, and engagement in higher-risk practices such as unsafe sexual activity or alcohol use, which may increase susceptibility to HBV infection. Additionally, men are less likely to participate in preventive health programs, including vaccination campaigns, potentially contributing to this disparity[34].

Not having a tattoo was associated with a lower risk of HBV infection. Individuals without tattoos were almost 0.3 times as likely to contract Hepatitis B compared to those with tattoos (Table 2.0). Tattooing, especially when performed under unregulated or unsafe conditions, can facilitate transmission through contaminated needles and equipment. Other studies have similarly reported associations between tattooing, body scarification, and HBV seropositivity [35–38]. Given the cultural and social significance of tattoos and related practices in many African communities, public health strategies should emphasize the importance of safe procedures and infection prevention practices in both formal and informal tattoo settings.

Finally, vaccination uptake was inversely associated with HBV infection, underscoring the protective role of full adherence to the hepatitis B vaccination schedule. When compared with those who have been vaccinated, those who have not been vaccinated against hepatitis B had 3.4-fold risk of getting infected with HBV. This finding supports the protective effect of the vaccine which has also been shown in Figure 1B, where there was a noticeable decreasing prevalence of HBV positivity with increasing number of vaccine shots[39]. Participants who did not complete the recommended three-dose series were more likely to be HBsAg-positive, in line with previous studies demonstrating that incomplete vaccination compromises long-term immunity[40,41]. In Ghana, despite the introduction of the HBV vaccine into the Expanded Program on Immunization (EPI) in 2002, adult coverage remains low, and poor compliance with vaccination schedules has been reported among healthcare workers and the general population[12,42]. Our findings highlight the urgent need to strengthen vaccination programs, ensure the availability and affordability of vaccines, and implement post-vaccination serological testing, particularly in high-risk occupational and informal sector groups. Adult catch-up vaccination campaigns, especially in informal sectors are recommended.

Although occupational category was not found to be a statistically significant predictor of HBV infection in this study, variations in prevalence were observed across different work groups. The highest prevalence was recorded among butchers and meat sellers (13.0%) and drivers/bus conductors (10.0%) [4], while relatively lower rates were seen among fishmongers (2.4%) and general goods storekeepers (2.3%). Tailors/seamstresses, market porters and food vendors exhibited intermediate prevalence levels of 8.2%, 7.7% and 7.6%, respectively. These findings, though not statistically significant, are noteworthy as they may reflect differences in occupational exposures and workplace practices. For instance, butchers and drivers may be at increased risk of accidental cuts, exposure to blood, or unregulated sharp object use, which are recognized routes of HBV transmission. By contrast, fishmongers and storekeepers may face fewer such exposures in the course of their work. The absence of statistical significance could be due to overlapping behavioral risk factors across occupations, relatively small subgroup sample sizes that limited statistical power, or the influence of protective factors such as vaccination in some occupational groups. Nonetheless, these occupational prevalence patterns provide useful insights into possible priority groups for HBV awareness, screening, and vaccination interventions within informal work settings.

Together, these findings have important public health implications. The persistence of intermediate-to-high HBV prevalence among market workers, a group that reflects the general adult population, suggests that Ghana remains far from achieving the WHO’s target of reducing hepatitis B incidence by 90% and mortality by 65% by 2030[29]. Targeted interventions such as increasing awareness of safe tattooing practices, promoting male-focused screening and vaccination campaigns, and improving compliance with vaccination schedules are essential steps toward reducing HBV burden in Ghana.

In contrast to findings from previous studies that have consistently identified blood transfusion, early sexual debut, multiple sexual partners, and household exposure to HBV-positive individuals as significant risk factors for HBV infection [21,33,43,44], our study did not demonstrate statistically significant associations for these variables. Several factors may explain these observations. First, the risk of transfusion-associated HBV transmission has declined substantially in Ghana following the introduction of routine HBV screening of donated blood, thereby reducing its contribution as a transmission route in contemporary settings [45,46]. Second, although sexual transmission remains an established pathway, our study population was largely older adults, many of whom may already have acquired immunity through prior vaccination, reducing the relative impact of sexual behaviors on current infection status. Additionally, the reliance on self-reported data for sexual history and family infection status may have introduced recall or social desirability bias, leading to underreporting and consequent dilution of associations. Finally, the occupational nature of the study population suggests that practices such as tattooing may play a more dominant role than traditional household or sexual risk factors, which could explain why these factors lost significance in multivariable models.

### Strengths and limitations

This study has some limitations. Being cross-sectional in nature, it cannot establish causality between the predictors and HBV infection. Additionally, recall bias and social desirability bias could affect the accuracy of self-reported data. Also, the use of rapid diagnostic tests (RDTs), while practical in field settings, may have lower sensitivity and specificity compared with ELISA or molecular assays. Nonetheless, the findings provide valuable insight into HBV epidemiology in an important urban working population and contribute to the evidence base guiding HBV prevention and control strategies in Ghana. The findings provide evidence to guide targeted interventions in informal sector populations, which are often underrepresented in surveillance data. Also, randomized sampling ensures the generalizability of our findings.

## Conclusion

Hepatitis B virus infection remains a significant public health challenge among Kejetia Market workers, with a prevalence of 7.36%, reflecting intermediate-to-high endemicity. Female gender, absence of tattooing, and getting vaccinated were associated lower risk of HBV infection. Strengthening vaccination uptake, promoting safe tattoo practices, and implementing targeted male-focused interventions are critical to achieving Ghana’s HBV elimination goals.

## Data Availability

The data that support the findings of this study are openly available in Dryad with DOI as 10.5061/dryad.cjsxksnkd

https://doi.org/10.5061/dryad.cjsxksnkd

## Acknowledgements

We are very much grateful to the management of Kejetia Market for their support during this study. We also express our deep appreciation to all individuals who willingly participated in this study.

## References

[1] Hepatitis B n.d. https://www.who.int/news-room/fact-sheets/detail/hepatitis-b (accessed August 31, 2025).

[2] Hsu YC, Huang DQ, Nguyen MH. Global burden of hepatitis B virus: current status, missed opportunities and a call for action. Nat Rev Gastroenterol Hepatol 2023 208 2023;20:524–37. 10.1038/s41575-023-00760-9.

[3] Stanaway JD, Flaxman AD, Naghavi M, Fitzmaurice C, Vos T, Abubakar I, et al. The global burden of viral hepatitis from 1990 to 2013: findings from the Global Burden of Disease Study 2013. Lancet 2016;388:1081–8. 10.1016/S0140-6736(16)30579-7.

[4] Abesig J, Chen Y, Wang H, Mwekele F, Irene S, Wu XY. Prevalence of viral hepatitis B in Ghana between 2015 and 2019: A systematic review and meta-analysis. PLoS One 2020;15. 10.1371/journal.pone.0234348.

[5] Dodoo ANO, Renner L, Van Grootheest AC, Labadie J, Antwi-Agyei KO, Hayibor S, et al. Safety monitoring of a new pentavalent vaccine in the expanded programme on immunisation in Ghana. Drug Saf 2007;30:347–56. 10.2165/00002018-200730040-00007/METRICS.

[6] Fofana DB, Somboro AM, Maiga M, Kampo MI, Diakité B, Cissoko Y, et al. Hepatitis B Virus in West African Children: Systematic Review and Meta-Analysis of HIV and Other Factors Associated with Hepatitis B Infection. Int J Environ Res Public Health 2023;20:4142. 10.3390/IJERPH20054142/S1.

[7] Efua SDV, Adwoa WD, Armah D. Seroprevalence of Hepatitis B virus infection and associated factors among health care workers in Southern Ghana. IJID Reg 2023;6:84–9. 10.1016/j.ijregi.2023.01.009.

[8] Nartey YA, Okine R, Seake-Kwawu A, Ghartey G, Asamoah YK, Senya K, et al. A nationwide cross-sectional review of in-hospital hepatitis B virus testing and disease burden estimation in Ghana, 2016 - 2021. BMC Public Health 2022;22. 10.1186/s12889-022-14618-3.

[9] Nyambah PK, Agjei R, Sarfo B. Seroprevalence and factors associated with Hepatitis B virus infection among students in two senior high schools in the Krachi Nchumuru district in Ghana-A cross-sectional study. BMC Res Notes 2023;16:1–11. 10.1186/S13104-023-06624-4/TABLES/4.

[10] Ofori-Asenso R, Agyeman AA. Hepatitis B in Ghana: A systematic review & meta-analysis of prevalence studies (1995-2015). BMC Infect Dis 2016;16:1–15. 10.1186/S12879-016-1467-5/FIGURES/9.

[11] Schulte PA, Chun HK. Climate Change and Occupational Safety and Health: Establishing a Preliminary Framework. J Occup Environ Hyg 2009;6:542–54. 10.1080/15459620903066008.

[12] Okwan DK, Scott GY, Takyi P, Boateng CO, Antwi PB, Abrampah AA, et al. A Multicentre Cross-Sectional Study on Hepatitis B Vaccination Coverage and Associated Factors Among Personnel Working in Health Facilities in Kumasi, Ghana. Biomed Res Int 2024;2024. 10.1155/2024/8899638.

[13] Mensah F, Shi G, Yu Q, Boadi EB, Andam FA, Bofah NA. The Impact of Resettlement in Urban Market Redevelopment on Income Inequality, Its Determinants, and Implications for the Resettled Population: Applying the Kejetia New Market Exemplar, Ghana. Sustain 2022;14:16682. 10.3390/SU142416682/S1.

[14] Okoye V. Street Vendor Exclusion in “Modern” Market Planning : A Case Study from Kumasi, Ghana. 2020.

[15] Goals SD. Epidemiology - 2016 - WHO - Draft global health sector strategy on viral hepatitis 2021:2016–21.

[16] Raosoft. Sample Size Calculator by Raosoft, Inc. n.d. http://www.raosoft.com/samplesize.html (accessed September 4, 2025).

[17] Amini A, Varsaneux O, Kelly H, Tang W, Chen W, Boeras DI, et al. Diagnostic accuracy of tests to detect hepatitis B surface antigen: A systematic review of the literature and meta-analysis. BMC Infect Dis 2017;17. 10.1186/s12879-017-2772-3.

[18] Nelson NP, Easterbrook PJ, McMahon BJ. Epidemiology of Hepatitis B Virus Infection and Impact of Vaccination on Disease. Clin Liver Dis 2016;20:607. 10.1016/J.CLD.2016.06.006.

[19] Osei E, Lokpo SY, Agboli E. Sero-prevalence of hepatitis B infection among blood donors in a secondary care hospital, Ghana (2014): a retrospective analysis. BMC Res Notes 2017;10:391. 10.1186/S13104-017-2733-3/TABLES/3.

[20] Mutaru A-M, Ahmed ·, Bawah T, Martin ·, Adokiya N. Socio-demographic determinants of hepatitis B virus seropositivity among adults in Tolon, Ghana: a community-based survey. Discov Viruses 2025 21 2025;2:1–9. 10.1007/S44370-025-00016-X.

[21] Jaldo MM, Joffe MWY, Zemedkun ES. Prevalence of hepatitis B virus and associated factors among blood donors in Hossana blood bank catchment area, Southern Ethiopia. BMC Infect Dis 2025;25:1–7. 10.1186/S12879-025-10550-0/TABLES/5.

[22] Nartey YA, Okine R, Seake-Kwawu A, Ghartey G, Asamoah YK, Senya K, et al. A nationwide cross-sectional review of in-hospital hepatitis B virus testing and disease burden estimation in Ghana, 2016 - 2021. BMC Public Health 2022;22:1–15. 10.1186/S12889-022-14618-3/TABLES/7.

[23] Helegbe GK, Tanko F, Aryee PA, Lotsu SA, Asaarik MJA, Anaba F. High Hepatitis B Seroprevalence, Low Knowledge, and Poor Attitude towards Hepatitis B Virus Infection among Market Women in Bolgatanga Metropolis in the Upper East Region of Ghana. J Trop Med 2020;2020. 10.1155/2020/4219413.

[24] Boachie J, Pidah D, Eshun H, Jingbeja E, Adjei PF, Adu P. Prevalence of Hepatitis B Viral Infection in Pregnant Women at the Suhum Municipality, Ghana. J Pregnancy 2024;2024:9438762. 10.1155/2024/9438762.

[25] Luuse A, Dassah S, Lokpo S, Ameke L, Noagbe M, Adatara P, et al. Sero-Prevalence of Hepatitis B Surface Antigen Amongst Pregnant Women Attending an Antenatal Clinic, Volta Region, Ghana. J Public Health Africa 2017;7:584. 10.4081/JPHIA.2016.584.

[26] Quintas AE, Cuboia N, Cordeiro L, Sarmento A, Azevedo L. Seroprevalence of Hepatitis B virus surface antigen among African blood donors: a systematic review and meta-analysis. Front Public Heal 2024;12:1434816. 10.3389/FPUBH.2024.1434816/FULL.

[27] Wondmeneh TG, Mekonnen AT. Epidemiology of hepatitis B virus infection among pregnant women in Africa: a systematic review and meta-analysis. BMC Infect Dis 2024;24:1–26. 10.1186/S12879-024-09839-3/METRICS.

[28] Razavi-Shearer DM, Gamkrelidze I, Pan CQ, Jia J, Berg T, Gray RT, et al. Global prevalence, cascade of care, and prophylaxis coverage of hepatitis B in 2022: a modelling study. Lancet Gastroenterol Hepatol 2023;8:879–907. 10.1016/S2468-1253(23)00197-8.

[29] WHO. Elimination of hepatitis by 2030 n.d. https://www.who.int/health-topics/hepatitis/elimination-of-hepatitis-by-2030?utm_source=chatgpt.com#tab=tab_1 (accessed September 4, 2025).

[30] Drazilova S, Kristian P, Janicko M, Halanova M, Safcak D, Dorcakova PD, et al. What is the role of the horizontal transmission of hepatitis b virus infection in young adult and middle-aged roma population living in the settlements in east Slovakia? Int J Environ Res Public Health 2020;17. 10.3390/IJERPH17093293,.

[31] Kafeero HM, Ndagire D, Ocama P, Kudamba A, Walusansa A, Sendagire H. Prevalence and predictors of hepatitis B virus (HBV) infection in east Africa: evidence from a systematic review and meta-analysis of epidemiological studies published from 2005 to 2020. Arch Public Heal 2021;79:1–19. 10.1186/s13690-021-00686-1.

[32] Kumasi G, Amidu N, Alhassan A, Obirikorang C, Feglo P, Majeed SF, et al. Sero-prevalence of hepatitis B surface (HBsAg) antigen in three densely populated communities in Kumasi, Ghana. J Med Biomed Sci 2012;1:59–65.

[33] Kafeero HM, Ndagire D, Ocama P, Kudamba A, Walusansa A, Sendagire H. Prevalence and predictors of hepatitis B virus (HBV) infection in east Africa: evidence from a systematic review and meta-analysis of epidemiological studies published from 2005 to 2020. Arch Public Heal 2021;79:1–19. 10.1186/S13690-021-00686-1/FIGURES/8.

[34] Rata Mohan DS, Jawahir S, Manual A, Abdul Mutalib NE, Mohd Noh SN, Ab Rahim I, et al. Gender differences in health-seeking behaviour: insights from the National Health and Morbidity Survey 2019. BMC Health Serv Res 2025;25:1–13. 10.1186/S12913-025-13020-0/TABLES/4.

[35] Belay AS, Abateneh DD, Yehualashet SS, Kebede KM. Hepatitis B virus infection and associated factors among adults in Southwest Ethiopia: Community-based cross-sectional study. Int J Gen Med 2020;13:323–32. 10.2147/IJGM.S259375;PAGE:STRING:ARTICLE/CHAPTER.

[36] Jafari S, Buxton JA, Afshar K, Copes R, Baharlou S. Tattooing and Risk of Hepatitis B: A Systematic Review and Meta-analysis. Can J Public Heal 2012;103:207–12. 10.1007/BF03403814/METRICS.

[37] Raslan E, AbdAllah M, Soliman S. The prevalence and determinants of hepatitis B among Egyptian adults: a further analysis of a country-representative survey. Egypt Liver J 2022;12. 10.1186/s43066-022-00207-x.

[38] Ko, Y. C., Lan, S. J., & Chang PY. An increased risk of hepatitis B virus infection from tattooing in Taiwan. - Abstract - Europe PMC n.d. https://europepmc.org/article/med/2362302 (accessed September 4, 2025).

[39] Bilounga Ndongo C, Eteki L, Siedner M, Mbaye R, Chen J, Ntone R, et al. Prevalence and vaccination coverage of Hepatitis B among healthcare workers in Cameroon: A national seroprevalence survey. J Viral Hepat 2018;25:1582–7. 10.1111/JVH.12974,.

[40] Obeng MA, Okwan DK, Adankwah E, Owusu PK, Gyamerah SA, Duah KB, et al. Seroconversion and Prevalence of Hepatitis B Surface Antigen among Vaccinated Health Care Workers in Ashanti Region, Ghana. Adv Med 2023;2023:1–8. 10.1155/2023/2487837.

[41] Guo Y, Gao P, Wang H, Wu J, Bai Q, Huang L, et al. Risk factors of hepatitis B virus infection between vaccinated and unvaccinated groups among spouses in 2006 and 2014: a cross-sectional study in Beijing. Hum Vaccines Immunother 2020;16:148–57. 10.1080/21645515.2019.1640428;WGROUP:STRING:PUBLICATION.

[42] Dassah S, Sakyi SA, Frempong MT, Luuse AT, Ephraim RKD, Anto EO, et al. Seroconversion of hepatitis B vaccine in young children in the Kassena Nankana District of Ghana: A cross-sectional study. PLoS One 2015;10. 10.1371/journal.pone.0145209.

[43] Opare-Asamoah K, Majeed SF, Owusu AO, Keelson KO, Owusu EA, Wondoh PM, et al. The Prevalence and Risk Factors of Hepatitis B Virus Infection Among Dwellers in A Peri-Urban District of Ghana: A Cross-Sectional Study. J Med Biomed Sci 2021;8:12–20. 10.54106/215.jmbs7z.

[44] Gedefaw G, Waltengus F, Akililu A, Gelaye K. Risk factors associated with hepatitis B virus infection among pregnant women attending antenatal clinic at Felegehiwot referral hospital, Northwest Ethiopia, 2018: An institution based cross sectional study. BMC Res Notes 2019;12:1–7. 10.1186/S13104-019-4561-0/TABLES/3.

[45] Walana W, Vicar EK, Kuugbee ED, Dari I, Bichenlib G, Aneba CN, et al. Transfusion transmissible infections among blood donors in Ghana: A 3-year multicentered health facility-based retrospective study. Heal Sci Reports 2023;6. 10.1002/HSR2.1681.

[46] Hadfield PY, Vechey GA, Bansah E, Nyahe M, Khuzwayo N, Tarkang EE. Transfusion-Transmissible Infections Among Blood Donors in a Regional Hospital in Ghana: A 6-Year Trend Analysis (2017-2022). J Int Assoc Provid AIDS Care 2024;23. 10.1177/23259582241274305.

